# E-cigarette Duration and Incident COPD Among Adults Aged 40 Years and Older with a Smoking History

**DOI:** 10.64898/2026.02.04.26345592

**Authors:** S. Cook, A.F. Brouwer, J.M.G. Taylor, KM. Cummings, D.A. Arenberg, N.L. Fleischer, R. Meza

## Abstract

**Importance:** Chronic obstructive pulmonary disease (COPD) remains a leading cause of morbidity and mortality in the United States (US), largely driven by cigarette smoking and characterized by progressive lung injury. While e-cigarettes are promoted as a less harmful alternative to cigarette smoking, their long-term health effects, including the impact of prolonged use on COPD incidence among adults who have smoked, are not well understood.

**Objective:** To evaluate the prospective association between duration of e-cigarette use and incident COPD among US adults aged 40 years or older with a history of cigarette smoking, and to determine whether baseline respiratory symptoms modify this association.

**Design, Settings, and Participants:** We used data from Waves 4 to 7 (2017-2022) of the Population Assessment of Tobacco and Health (PATH) Study, a nationally representative US longitudinal cohort study. Our analysis included adults aged 40 years or older who currently or formerly smoked cigarettes.

**Main Outcomes and Measures:** The outcome was incident, self-reported COPD diagnosis. The main exposure was the time-varying duration of e-cigarette use. Baseline functionally important respiratory symptoms were defined by a validated index. Multivariable models adjusted for demographics, COPD risk factors, and detailed tobacco use history, including cigarette smoking status, time since quitting, and pack-years.

**Results:** Among 4,895 adults aged 40 year or older who currently or formerly smoked cigarettes, 408 reported an incident COPD diagnosis. Among individuals with baseline respiratory symptoms, longer e-cigarette use duration was associated with increased COPD risk (adjusted hazard ratio [AHR]: 1.28, 95% CI: 1.16, 1.40), whereas no significant association was observed among those without baseline respiratory symptoms (AHR: 1.01, 95% CI: 0.92, 1.12). Results were consistent after adjusting for cumulative cigarette exposure and other risk factors and remained robust across multiple sensitivity analyses.

**Conclusion and Relevance:** Prolonged e-cigarette use may increase COPD risk among individuals with pre-existing respiratory vulnerabilities. Although switching from combustible cigarettes remains an important harm reduction strategy, behavioral counseling and pharmacotherapy should be prioritized for those at high risk for COPD, with e-cigarette cessation support available to high-risk former smokers. Continued surveillance and research are warranted as e-cigarette products and use patterns evolve.

**Key Points:** *Question:* Does longer e-cigarette use increase COPD risk in adults with a smoking history?

*Findings:* In this national cohort study of U.S. adults aged 40+ who currently or formerly smoked, e-cigarette duration was associated with higher self-reported COPD incidence among individuals with respiratory symptoms at baseline (adjusted hazard ratio [AHR] 1.28, 95% CI 1.16, 1.40) but not among those without symptoms (AHR 1.01, 95% CI 0.92, 1.12).

*Meaning:* Prolonged e-cigarette use may increase COPD risk among individuals with respiratory vulnerabilities. While cigarette cessation should remain the priority, evidence-based e-cigarette cessation strategies are needed to prevent long-term use in this population.

## Introduction

Chronic obstructive pulmonary disease (COPD) is one of the leading causes of mortality in the United States (US),^1,2^ expected to account for the loss of more than 45 million quality-adjusted life-years from 2019 to 2038.^3^ COPD is a slowly developing and progressive respiratory disease that is characterized by reduced airflow and increased pulmonary inflammation.^4,5^ It is often caused by high levels of exposure to noxious particles or gases,^4,5^ and, in particular, exposure to cigarette smoke is a major contributor to COPD development.^5-8^ Adults with a long duration of cigarette use^9^ or a large cumulative exposure (cigarette pack-years ≥20^7^) are at particularly high risk for developing COPD.

E-cigarettes are alternative nicotine-delivery systems that vaporize nicotine-containing liquids rather than combust tobacco. While e-cigarette use is likely substantially less harmful to health than cigarette smoking,^10,11^ it is currently unknown whether and to what extent exposure to e-cigarettes may increase COPD risk. Evidence from clinical studies suggests that e-cigarettes may induce acute inflammatory responses and immune dysregulation,^12-17^ and several cross-sectional epidemiological studies have associated e-cigarette use with increased respiratory disease^18-20^ and COPD prevalence.^21-24^ However, the validity of these cross-sectional associations has been questioned,^25-29^ and, among the many methodological limitations identified in studies attempting to quantify the harmful health effects of e-cigarette use,^30^ reverse causation concerns and inadequate adjustment for cigarette smoking histories are among the most serious. Specifically, because most adults aged 40 and over who use e-cigarettes have extensive histories of cigarette smoking,^25,30-32^ with many making the switch only after developing health issues (including early signs of COPD),^30,33^ prospective longitudinal studies that carefully account for both cigarette and e-cigarette use histories are needed to help mitigate confounding and measurement concerns. Such studies are starting to emerge,^25,34,35^ and in a longitudinal study using data from the first 5 waves of data from the Population Assessment of Tobacco Health (PATH) Study (2013–19), we found that current e-cigarette use was not associated with an increased risk of COPD incidence after accounting for current smoking status and cigarette pack-years.^25^ However, this previous study was limited by the relatively short duration of e-cigarette exposure.

To date, no studies have examined whether duration of e-cigarette use, not just current use, is associated with COPD risk among those who have ever smoked. In cigarette research, we know that longer durations of smoking are strongly predictive of COPD,^8,9,36^ marking duration as a key factor in the physiological development of chronic lung disease.^9^ Similarly, prolonged and regular use of e-cigarettes might also be expected to increase COPD risk by causing cumulative inflammation and airflow obstruction.^14^ However, because e-cigarette products are relatively new, with little adoption prior to 2012,^37^ cumulative e-cigarette exposure is generally much lower compared to cigarettes, measured in years, not decades. Moreover, almost all adults aged 40 and older who use e-cigarettes either currently or formerly smoked cigarettes,^28-30^ meaning they may have already accumulated significant risk for COPD from cigarette smoking prior to e-cigarette use initiation. It is thus important to better understand whether even relatively short durations of e-cigarette use duration may further increase COPD risk in individuals already at high risk.

In this study, we assessed associations between e-cigarette use duration and self-reported incident COPD in a nationally representative, prospective longitudinal cohort study of US adults who ever smoked, adjusting for cigarette smoking history and other potential confounders. The cohort was followed for approximately five years to-date (2017–22), with e-cigarette and cigarette use information collected at each wave, allowing us to assess the impact of time-varying e-cigarette duration alongside changes in cigarette smoking. Given that e-cigarettes are promoted as a harm-reducing alternative to cigarettes,^38^ potentially leading adults who smoke or recently quit to switch to e-cigarettes after experiencing respiratory symptoms, we further examined whether functionally important respiratory symptoms (FIRS)^39^ at baseline moderated the association between e-cigarette duration and COPD risk.

## Methods

### Data

We analyzed restricted-use data from adults aged 40 and older who ever smoked cigarettes at least 100 cigarettes in their lifetime in Waves 4–7 (2017–22) of the Population Assessment of Tobacco and Health (PATH) Study, a nationally representative longitudinal study of the non-institutionalized civilian US population. Adults with no previous COPD diagnosis at Wave 4 who completed at least one follow-up were included and were followed across Waves 5–7 for self-reported incident COPD. This age threshold is consistent with national COPD cohort studies^40^ and reflects the rarity of COPD diagnosis before age 40.^41^ Participants contributed data until reporting COPD or until their last observation. Additional details on the PATH Study, including survey design^42,43^ and restricted-use data access^44^ are available elsewhere, and details about the construction of the analytic sample are provided in Appendix Figure 1. This study was approved by the institutional review board at the University of Michigan (HUM00162265) and followed STROBE reporting guidelines.

### Measures

At each follow-up interview, incident COPD was assessed via self-report. Participants were asked: “In the past 12 months, has a doctor, nurse, or other health professional told you that you had… (1) COPD, (2) chronic bronchitis, or (3) emphysema?” In accordance with clinical criteria, anyone reporting a new diagnosis of any of these conditions was classified as having incident COPD.

PATH study participants who reported ever ‘fairly regular’ e-cigarette use reported their ages of initiation and, if applicable, quitting. E-cigarette duration was calculated as the difference between current age and age at initiation (current use) or age at quit (former use) at each wave. Participants who never used e-cigarettes at least ‘fairly regularly’ were assigned a duration value of zero years. E-cigarette duration was included as a time-varying covariate, lagged by one wave (t-1) to ensure the duration exposure preceded the COPD diagnosis.

Baseline sociodemographic covariates included age (continuous), sex (male, female), race and ethnicity (Hispanic, Non-Hispanic (NH) White, NH Black, Another race or ethnicity), education (high school or less, some college, bachelor’s degree or more), and health insurance status (any, none). Additional COPD risk factors included obesity (body mass index (BMI) >=30) and second-hand smoke exposure (hours of ‘close’ exposure during the past 7 days, range 0-100); second-hand smoke exposure was included as a time-varying covariate (t-1).

Baseline respiratory symptom burden was assessed using the Functionally Important Respiratory Symptoms (FIRS) index, a validated, self-reported measured in the PATH dataset that evaluates the frequency and severity of symptoms such as cough, breathlessness, and wheeze.^45^ Consistent with prior validation research and published recommendations,^45^ we classified participants with a FIRS score of 3 or higher (FIRS 3+) as having significant respiratory symptom burden for our primary analysis. The cutoff of 3 has been empirically derived and validated to distinguish clinically meaningful levels of respiratory symptoms.

To address tobacco-related confounding, we included three time-varying (t-1) tobacco use variables: (1) cigarette smoking status (current, former: quit < 2 years, 2-6 years, >6 years), (2) established use of other combustible tobacco products (traditional cigars, filtered cigars, cigarillos, hookah, and pipes); and (3) cigarette pack years, calculated by multiplying years of cigarette smoking by the average number of packs per day while individuals smoked.

### Statistical analysis

We calculated weighted descriptive statistics for sociodemographic characteristics and smoking behaviors overall and by COPD outcome. We then compared cigarette smoking histories by e-cigarette duration status across exposure waves (Waves 4–6). To assess the association between e-cigarette use duration and incident self-reported COPD at follow-up (Waves 5–7) among adults aged 40+ who ever smoked cigarettes in the Wave 4 cohort (person n =4,859; risk-period n =11,941), we fitted discrete-time survival models following established methodologies.^25,34,46^ To evaluate whether this association varied by baseline respiratory risk, we included an interaction term and stratified our models by functionally important respiratory symptoms (FIRS 3+) at baseline. This approach allowed us to examine potential heterogeneity in effects across risk strata. Hazard ratios (HRs) and 95% confidence intervals (CIs) were estimated using a general linear model with a binomial distribution and a complementary log–log link, and Wave 4 weights were used to ensure national representativeness at baseline.^43^

We conducted several sensitivity analyses to test the robustness of our findings. First, we considered whether survey weighting impacted the results by using the longitudinal weights, which excluded participants who did not complete surveys in every wave. Second, because young adults are more likely to use e-cigarettes,^37,47^ and because surveys have found the COPD is occasionally self-reported before age 40,^48^ we expanded our analysis to include adult participants aged 25 years and older. Third, to assess the potential impact of adjusting for baseline respiratory risk, we conducted a sensitivity analysis where FIRS 3+ was removed as a covariate to examine whether adjusting for it as a confounder introduced bias due to time-dependent confounding and collider stratification.^49,50^ Fourth, because the health effects of e-cigarette use may be greatest among adults who continue to smoke cigarettes,^51^ we estimated an interaction between e-cigarette duration and smoking status (current vs. former) to assess whether the association with incident COPD differed by time-varying smoking status. we tested an interaction between e-cigarette duration and cigarette pack-years to assess whether the association between e-cigarette duration and incident COPD varied by cumulative smoking history. Finally, we evaluated the interaction between FIRS and e-cigarette duration using a lower threshold for respiratory symptoms (FIRS 2+) to examine sensitivity to a more inclusive respiratory symptom cut-off.

## Results

Among adults aged 40 years and older at baseline (n=4,895) there were 408 incident COPD cases across the follow-up period, representing an average weighted hazard rate of 1.4% per wave (Table 1; lifetable available in Appendix Table A1). Among participants diagnosed with COPD at follow-up, 58.2% currently and 42.8% formerly smoked cigarettes, with a with a mean of 32.9 cigarette pack years (SD=32.9). Additionally, 15.3% reported e-cigarette duration at baseline, with a mean duration of 2.5 years (SD=2.7). In contrast, among participants without a COPD diagnosis, 33.8% currently and 66.2% formerly smoked cigarettes (p<0.001) with a mean of 20.1 cigarette pack years (SD=22.3, p<0.001). For e-cigarette use, 8.4% reported at least one year of e-cigarette duration at baseline, with a mean duration of 2.4 years (SD=2.4, p<0.001). Baseline respiratory symptoms (FIRS 3+) were also more common among those with a COPD diagnosis at follow-up 34.3% compared to those with no COPD diagnosis 13.9% (p<0.001). Furthermore, participants with at least one year of e-cigarette duration had a higher prevalence of respiratory symptoms and elevated FIRS scores at baseline compared to those with no e-cigarette use duration (Appendix Table A2).

**Table 1.**
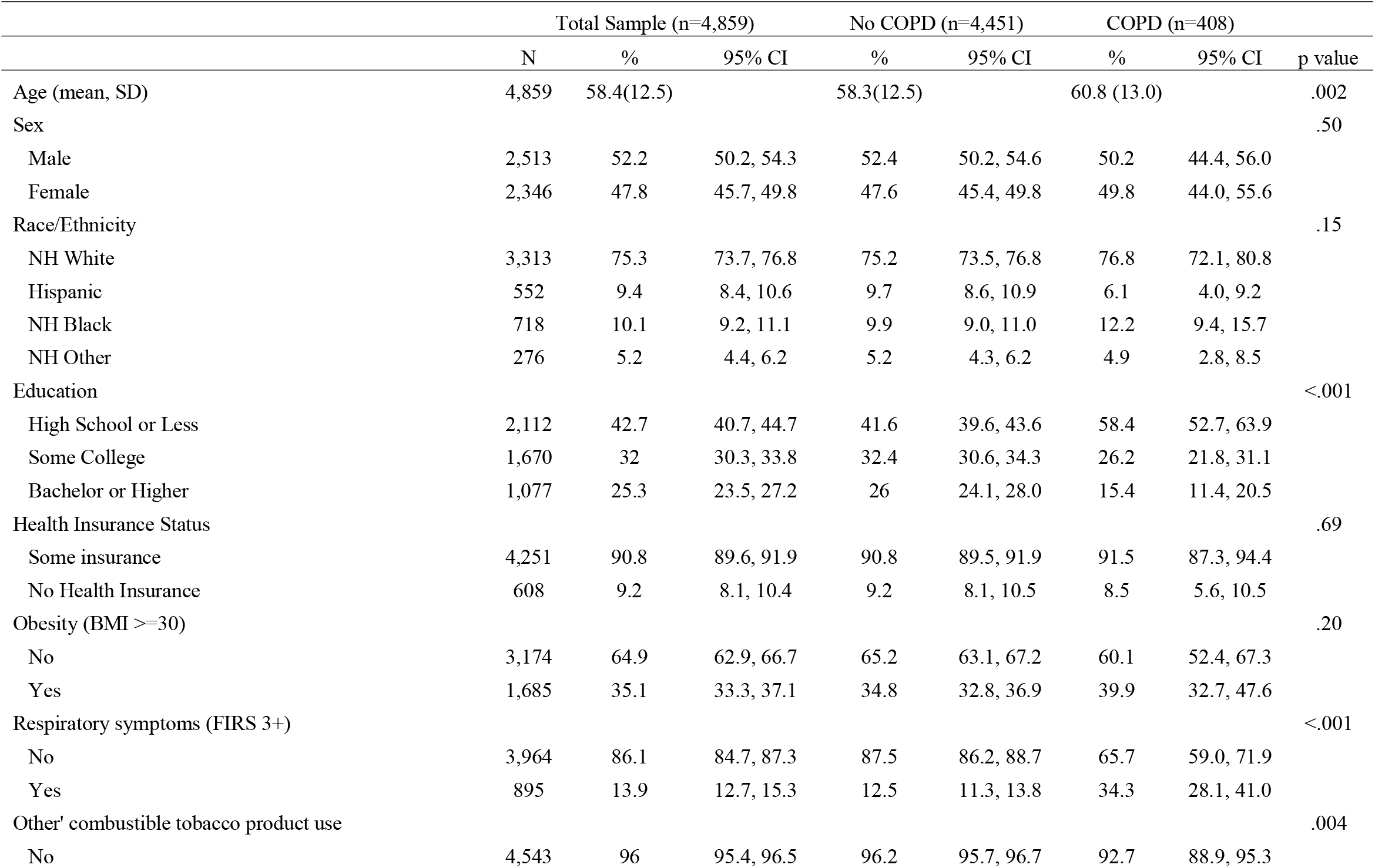
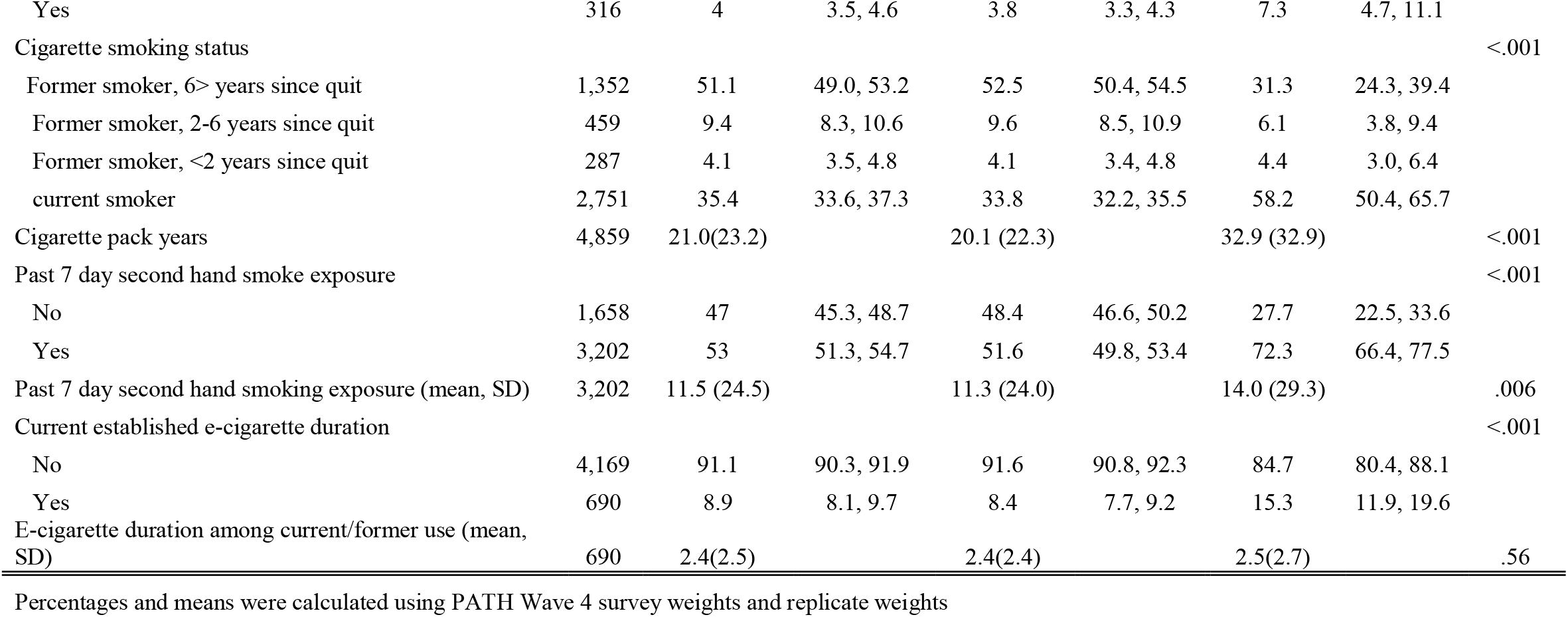
Baseline characteristics by incident COPD outcome among ever-smoking adults 40+.

At each wave, more than half of adults who reported any e-cigarette duration also currently smoked cigarettes (53.0%–64.8% across waves, Table 2), compared to less than one third of adults without e-cigarette use duration (24.6%–32.6%, p<0.001). At baseline, adults with at least one year of e-cigarette use duration had higher cigarette pack-years (mean=26.0, SD=29.5) and more hours of second-hand smoke exposure (mean=15.3, SD=31.4) compared to adults with no e-cigarette use (CPY mean=20.4, SD=22.5, p<0.001; second-hand smoke exposure hours mean=11.0, SD=23.3, p<0.001).

**Table 2.** Tobacco-related covariates by e-cigarette duration among ever-smoking adults 40+.

|  | Wave 4 |  |  |  |  | Wave 5 |  |  |  |  |
| --- | --- | --- | --- | --- | --- | --- | --- | --- | --- | --- |
|  | at least one year of e-cigarette use |  |  |  |  | at least one year of e-cigarette use |  |  |  |  |
|  | No |  | Yes |  | P<br>value | No |  | Yes |  | P<br>value |
|  | % | 95% CI | % | 95% CI |  | % | 95% CI | % | 95% CI |  |
| Smoking Status |  |  |  |  | <.001 |  |  |  |  | <.001 |
| Former smoking, 6> years since quit | 55.5 | 53.4,<br>57.7 | 5.2 | 3.1, 8.6 |  | 59.4 | 57.2,<br>61.6 | 12.4 | 8.1, 18.4 |  |
| Former smoking, 2-6 years since quit | 8.4 | 7.4, 9.7 | 19.2 | 15.8,<br>23.1 |  | 8.8 | 7.7, 10.0 | 18.6 | 14.9,<br>22.9 |  |
| Former smoking, <2 years since quit | 3.4 | 2.9, 4.2 | 10.8 | 8.2, 14.2 |  | 3.2 | 2.5, 4.2 | 8.6 | 6.0, 12.3 |  |
| current smoking | 32.6 | 30.7,<br>34.9 | 64.8 | 60.6,<br>68.9 |  | 28.6 | 26.9,<br>30.3 | 60.4 | 55.3,<br>65.2 |  |
| Cigarette pack-years (mean, SD) | 20.5<br>(22.5) |  | 26.0<br>(29.5) |  | <.001 | 19.6<br>(21.5) |  | 26.4<br>(29.3) |  | <.001 |
| Other combustible product use |  |  |  |  | <.001 |  |  |  |  | .013 |
| No | 96.3 | 95.7,<br>96.8 | 92.8 | 90.6,<br>94.5 |  | 96 | 95.3,<br>96.6 | 94 | 91.5,<br>95.5 |  |
| Yes | 3.7 | 3.2, 4.3 | 7.2 | 5.5, 9.4 |  | 4 | 3.4, 4.7 | 6 | 4.5, 8.5 |  |
| second-hand smoke exposure |  |  |  |  | <.001 |  |  |  |  | <.001 |
| No | 49.3 | 47.7,<br>51.1 | 23.7 | 20.1,<br>27.8 |  | 53.2 | 50.6,<br>55.7 | 24.5 | 20.3,<br>29.2 |  |
| Yes | 50.7 | 48.9,<br>52.5 | 76.3 | 72.2,<br>79.9 |  | 46.8 | 44.3,<br>49.4 | 75.6 | 70.8,<br>79.7 |  |
| second-hand hours of exposure (mean, SD) | 11.0<br>(23.3) |  | 15.3<br>(31.9) |  | <.001 | 9.9 (21.7) |  | 14.7<br>(32.7) |  | <.001 |

Table 2. Tobacco-related covariates by e-cigarette duration among ever-smoking adults 40+, continued
|  | Wave 6 |  |  |  | p<br>value |
| --- | --- | --- | --- | --- | --- |
|  | at least one year of e-cigarette use |  |  |  |  |
|  | No |  | Yes |  |  |
|  | % | 95% CI | % | 95% CI |  |
| Smoking Status |  |  |  |  | <.001 |
| Former smoking, 6+ years since quit | 67.5 | 65.2,<br>69.6 | 21.9 | 16.9,<br>27.9 |  |
| Former smoking, 2-6 years since quit | 4.7 | 3.9, 5.8 | 15.4 | 11.9,<br>19.7 |  |
| Former smoking, <2 years since quit | 3.2 | 2.6, 3.8 | 9.7 | 7.2, 13.0 |  |
| current smoking | 24.6 | 22.9,<br>26.4 | 53 | 47.6,<br>58.4 |  |
| Cigarette pack-years (mean, SD) | 19.2<br>(21.1) |  | 26.1<br>(29.0) |  | <.001 |
| Other combustible product use |  |  |  |  | <.001 |
| No | 97.1 | 96.5,<br>97.7 | 93.1 | 90.5,<br>95.0 |  |
| Yes | 2.9 | 2.3, 3.6 | 6.9 | 5.0, 9.5 |  |
| second-hand smoke exposure |  |  |  |  | <.001 |
| No | 60.5 | 58.2,<br>62.7 | 35 | 29.5,<br>41.0 |  |
| Yes | 39.5 | 37.2,<br>41.8 | 65 | 59.0,<br>70.5 |  |
| second-hand hours of exposure (mean, SD) | 11.4<br>(23.4) |  | 16.1<br>(35.8) |  | .005 |

All values were calculated using PATH Wave 4 survey weights and replicate weights

In an unadjusted model examining the prospective association between e-cigarette use duration and self-reported COPD incidence (Table 3), e-cigarette duration was associated with an increased hazard of COPD (Model 1, hazard ratio [HR] =1.15 per year of e-cigarette use, 95% CI 1.08, 1.21). E-cigarette duration remained significantly associated with increased COPD risk in all subsequent main effects models: adjusting for sociodemographic and COPD risk factors (Model 2, adjusted hazard ratio [AHR] = 1.15, 95% CI 1.09, 1.22); additionally adjusting for cigarette smoking status and other combustible tobacco product use (Model 3, AHR= 1.12, 95% CI 1.05, 1.19); and additionally adjusting for cigarette pack-years (Model 4, AHR= 1.10, 95% CI 1.04, 1.17).

**Table 3.** E-cigarette use duration and incident COPD among ever-smoking adults 40+.

|  | Model 1<sup>a</sup> |  | Model 2<sup>b</sup> |  | Model 3<sup>c</sup> |  | Model 4<sup>d</sup> |  |
| --- | --- | --- | --- | --- | --- | --- | --- | --- |
|  | Hazard | 95% CI | Hazard | 95% CI | Hazard | 95% CI | Hazard | 95% CI |
| Duration of e-cigarette use | 1.15 | 1.08, 1.21 | 1.15 | 1.09, 1.22 | 1.12 | 1.05, 1.19 | 1.1 | 1.04, 1.17 |
| Age |  |  | 1.03 | 1.02, 1.05 | 1.05 | 1.03, 1.06 | 1.04 | 1.03, 1.06 |
| Sex (Female=1) |  |  | 1.09 | 0.85, 1.43 | 1.1 | 0.85, 1.43 | 1.2 | 0.92, 1.56 |
| Race/Ethnicity |  |  |  |  |  |  |  |  |
| Hispanic |  |  | 0.69 | 0.43, 1.13 | 0.66 | 0.41, 1.05 | 0.86 | 0.54, 1.37 |
| White |  |  | REF | REF | REF | REF | REF | REF |
| Black |  |  | 1.15 | 0.83, 1.59 | 0.92 | 0.68, 1.26 | 1.05 | 0.78, 1.43 |
| Other |  |  | 1.13 | 0.63, 2.05 | 1.09 | 0.60, 1.98 | 1.19 | 0.66, 2.14 |
| Education |  |  |  |  |  |  |  |  |
| High School or Less |  |  | 2.21 | 1.46, 3.34 | 1.75 | 1.14, 2.70 | 1.5 | 0.99, 2.29 |
| Some College |  |  | 1.26 | 0.87, 1.86 | 1.11 | 0.75, 1.66 | 1.00 | 0.68, 1.48 |
| Bachelor Degree or Higher |  |  | REF | REF | REF | REF | REF | REF |
| Uninsured |  |  | 0.76 | 0.49, 1.18 | 0.73 | 0.48, 1.12 | 0.72 | 0.47, 1.11 |
| Obesity (BMI>30) |  |  | 1.15 | 0.81, 1.63 | 1.36 | 0.97, 1.90 | 1.32 | 0.95, 1.84 |
| FIRS 3+ |  |  | 3.43 | 2.53, 4.64 | 2.91 | 2.16, 3.92 | 2.67 | 1.95, 3.67 |
| Time-varying second-hand smoke exposure<sup>e</sup> |  |  | 1.1 | 1.05, 1.16 | 1.05 | 0.99, 1.10 | 1.04 | 0.98, 1.09 |
| Time-varying 'other' combustible tobacco use |  |  |  |  | 1.46 | 0.90, 2.37 | 1.54 | 0.96, 2.47 |
| Time-Varying Smoking Status Indicator |  |  |  |  |  |  |  |  |
| Former Smoker, 6 > years quit |  |  |  |  | 0.34 | 0.23, 0.49 | 0.37 | 0.25, 0.54 |
| Former smoker, 2-5 years quit |  |  |  |  | 0.4 | 0.24, 0.65 | 0.5 | 0.31, 0.79 |
| Former smoker, < 2 years quit |  |  |  |  | 1.09 | 0.70, 1.71 | 1.17 | 0.75, 1.84 |
| Current Smoker |  |  |  |  | REF | REF | REF | REF |
| Time-varying cigarette pack-years |  |  |  |  |  |  | 1.38 | 1.17, 1.64 |

Person-level analytic n= 4,859. Risk set analytic n=11,941. All analyses were weighted using Wave 4 survey weights and replicate weights
<sup>a</sup> e-cigarette exposure; <sup>b</sup> sociodemographic variables added; <sup>c</sup> smoking covariates added; <sup>d</sup> pack-years added; <sup>e</sup> variable rescaled to intervals of 10 hours

To assess whether baseline respiratory symptoms modified the association between e-cigarette use duration and incident COPD, we tested an interaction between e-cigarette duration and FIRS 3+ status at baseline (p=0.01; Appendix Table A3). As shown in the forest plot (Figure 1), e-cigarette duration was significantly associated with COPD risk in multivariable models among participants with baseline respiratory symptoms (FIRS 3+: AHR=1.28, 95% CI 1.16, 1.40) but not among those without symptoms (FIRS <3: AHR=1.01, 95% CI 0.92, 1.12). Full model estimates are provided in Appendix Table A4.

In the sensitivity analyses restricted to the longitudinal cohort who participated in all Waves 4-7 (Appendix Table A5), expanded to include adults aged 25 years and older (Appendix Table A6), and removing FIRS 3+ as a covariate (Appendix Table A7), the substantive results remained consistent with the main findings. Moreover, in sensitivity analyses assessing the moderating effects of current cigarette smoking status (p= 0.757; Appendix Table A8) and cigarette pack-years (p=0.659; Appendix Table A9), the association between e-cigarette duration and incident COPD did not significantly differ by these smoking-related variables. The interaction between e-cigarette duration and FIRS 2+ at baseline was not statistically significant (p= 0.077; Appendix Table A10), although the results from stratified models (Appendix Table A11) indicated that e-cigarette duration was associated with increased COPD risk for participants with FIRS 2+ at baseline (AHR=1.18, 95% CI 1.09, 1.29) but not for those without baseline symptoms (AHR=1.01, 95% CI 0.90, 1.13).

## Discussion

In this nationally representative cohort of US adults aged 40 and over with a history of cigarette smoking, we observed that the association between e-cigarette use duration and incident COPD varied according to baseline respiratory symptom severity. Specifically, longer e-cigarette use duration was associated with a higher hazard of self-reported incident COPD among participants with more severe baseline respiratory symptoms (FIRS 3+), while no significant association was observed among those below this threshold. To our knowledge, this is the first study to explore this association, and our results persisted in models adjusting for time-varying tobacco-related confounders, including cigarette pack-years, and across multiple sensitivity tests.

Cigarette smoking duration is a well-established predictor of COPD,^8,9,36^ and our findings suggest that longer durations of e-cigarette use may be associated with increased COPD risk among individuals with pre-existing respiratory symptoms. E-cigarette vapors often contain chemicals, such as acetaldehyde and formaldehyde,^16^ that can induce inflammatory responses, increase oxidative stress, and disrupt immune system functioning, although to a lesser extent than from cigarette smoke.^12-17^ Prolonged exposure to these toxicants, particularly exposure spanning multiple years, may further compromise respiratory health and increase COPD risk, with the effect appearing most pronounced among those with already diminished respiratory function. Because our sample included adults aged 40 years and over at baseline who currently or formerly smoked cigarettes,^25^ the impact of e-cigarette duration is likely cumulative, building on prior exposures and contributing to the natural history of COPD progression from cigarette smoking.^4^

Our findings suggest that many adults who used e-cigarettes for an extended duration may have already been on the path to developing COPD at the beginning of follow-up. Although we could not determine whether e-cigarette use preceded respiratory symptoms, e-cigarettes are often used as a smoking cessation aid,^52,53^with many adults switching after experiencing adverse health effects.^30,33^ Thus, prolonged e-cigarette exposure may exacerbate COPD risk in those already symptomatic. Importantly, prior cohort studies have demonstrated that respiratory symptoms frequently precede a formal COPD diagnosis and may represent an early or transitional phase of airway disease.^54,55^ This raises the possibility that individuals with significant symptom burden were already experiencing undiagnosed of early-stage COPD, even without conformation from spirometry testing. For adults with significant smoking histories and pre-existing symptoms, extended e-cigarette use may accelerate respiratory decline. Cigarette cessation should remain a priority for all smokers; however, e-cigarette cessation should also be encouraged once cigarette smoking cessation is achieved. Behavioural counselling and pharmacotherapy, both evidence-based methods for patients with respiratory diseases,^56^ may be preferable as long-term cessation strategies for this at-risk population.

Our study has several limitations. First, we did not develop a pack-year analogue or capture intensity of e-cigarette use, primarily because of the lack of standardized and validated metrics across diverse e-cigarette products.^57^ Product heterogeneity—such as device type, nicotine concentration, and pod size^57^—and the limited availability of product-specific data in epidemiological surveys make it difficult to accurately quantify intensity of e-cigarette exposure.^30^ Second, e-cigarette duration was based on current past 30-day use at each wave, which assumes continuous use throughout the year and may capture both intermittent and persistent use, potentially reducing analytic precision (as e-cigarette use tends to be more transitory than cigarette smoking).^58,59^ Third, our measure of duration did not account for changing product types as e-cigarettes continue to evolve, which may affect exposure assessments.^30^ Fourth, both e-cigarette duration and COPD diagnosis were self-reported, introducing potential measurement error. Notably, our COPD diagnoses were not confirmed with spirometry, the clinical gold standard. Recent analyses of NHANES data indicate that self-reported COPD underestimates prevalence compared to spirometry criteria,^60^ suggesting our study may underestimate the true burden of COPD. Future research should incorporate clinical measures to validate COPD status. Fifth, the study period overlapped with the EVALI outbreak and the emergence of COVID-19, which may have influenced respiratory diagnoses and healthcare utilization patterns. Sixth, residual confounding by baseline respiratory symptom severity may have influenced the association between prolonged e-cigarette use and COPD risk, even with stratification and interaction modeling. Further disentangling these pathways is an important task for future research. Seventh, our findings were based on approximately five years of longitudinal data, and the results should be updated as more cigarette-naïve adults who use e-cigarettes age into higher-risk categories for COPD. Finally, it is plausible that the observed association between e-cigarette duration and COPD may be influenced, in part, by increased surveillance and respiratory functioning testing among e-cigarette users, particularly if they were already experiencing respiratory symptoms or were impacted by concerns about e-cigarette-related lung injury which occurred during our analysis period.^61^ Future research should investigate patterns of respiratory testing by e-cigarette use status at the population level, to better distinguish the effects of e-cigarette use and duration from disease onset versus diagnosis.

## Conclusion

In this nationally representative cohort of U.S. adults aged 40 years or over who ever smoked cigarettes, we found that longer durations of e-cigarette use were associated with increased COPD incidence among those with respiratory symptoms at baseline. These findings suggest that although e-cigarettes may pose fewer risks than combustible cigarettes, prolonged e-cigarette use—especially among individuals with pre-existing respiratory vulnerabilities—could potentially contribute to continued disease progression. Our results do not change the fact that supporting individuals in transitioning away from combustible cigarettes remains a critical harm reduction strategy. Rather, they underscore the potential importance of prioritizing behavioural counselling and pharmacotherapy as first-line cigarette cessation strategies for those at risk of developing COPD. Furthermore, high-risk patients who have quit smoking with the aid of e-cigarettes may benefit from additional resources to support e-cigarette cessation. Ongoing surveillance and research are needed to further inform best practices for tobacco harm reduction as e-cigarette products and usage patterns evolve.

## Data Availability

The data used in this study are from the Population Assessment of Tobacco and Health (PATH) Study, which are restricted and not publicly available due to privacy and data use agreements. Interested researchers can apply for access to PATH restricted-use datasets through the National Addiction Data Archive Program (NAHDAP) at https://www.icpsr.umich.edu/web/NAHDAP/studies/36231

## Acknowledgments

Research reported in this paper were supported by the National Cancer Institute of the National Institutes of Health (NIH) and the FDA Center for Tobacco Products (CTP) under Award Number 2U54CA229974. The content is solely the responsibility of the authors and does not necessarily reflect the views of the NIH or FDA. The authors wish to acknowledge the participants and organizers of the Tobacco Centers for Regulatory Science (TCORS) annual meeting (2024; Bethesda, MD), whose discussion and input contributed significantly to the refinement of this paper.

## Funding

Research reported in this publication was supported by the National Cancer Institute of the National Institutes of Health (NIH) and FDA Center for Tobacco Products (CTP) under Award Number 2U54CA229974. The content is solely the responsibility of the authors and does not necessarily represent the official views of the NIH or the Food and Drug Administration.

## Conflict of interest declaration

Drs. Cook, Brouwer, Taylor, Arenberg, Fleischer, and Meza report no conflicts of interest. Dr. Cummings also has been a paid expert witness in litigation against the cigarette industry.

## Notes

### Author Declarations

Ethics committee/IRB of University of Michigan gave ethical approval for this work.

