## Supplemental Files for "E-cigarette Duration and Incident COPD Among Adults Aged 40 Years and Older with a Smoking History"

Supplemental Material

Figure A1. Flowchart of sample selection for analytic sample

8,340 adult respondents aged 40+ participated in the W4 survey with established smoking history

1,819 respondents had a COPD outcome (COPD, chronic bronchitis, emphysema) at W4

6,521 adult respondents aged 40+ at baseline without a baseline COPD outcome

81

749 baseline respondents did not respond to any of the follow-up waves (W5-W7)

5,772 adult respondents aged 40+ did not have baseline COPD and completed at least one follow-up wave

634 respondents participated in at least one follow-up interview but did not have any follow-up information on COPD outcomes

5,138 adult respondents aged 40+ at baseline participated at follow-up and reported information about COPD

279 respondents (5.4%) had missing data on independent variable(s)

4,859 adult respondents aged 40+ at baseline participated at follow-up and had complete information on independent variables

Table A1. Life tables for incident self-reported COPD among ever-smoking adults 40+

| Interval | Total | COPD Diagnosis | Censored | Hazard Estimate^1^ |
| --- | --- | --- | --- | --- |
| Period 1 (W4-W5) | 4,859 | 198 | 758 | 0.0313 |
| Period 2 (W5-W6) | 3,903 | 139 | 585 | 0.0285 |
| Period 3 (W6-W7) | 3,179 | 71 | 3,108 | 0.0167 |
| Totals | 11,941 | 408 |  | 0.0144 |
| ^1^ Hazard estimates were calculated using Wave 4 survey weights and replicate weights | | | | |

Table A2. Functionally important respiratory symptoms by e-cigarette use duration among ever-smoking adults 40+

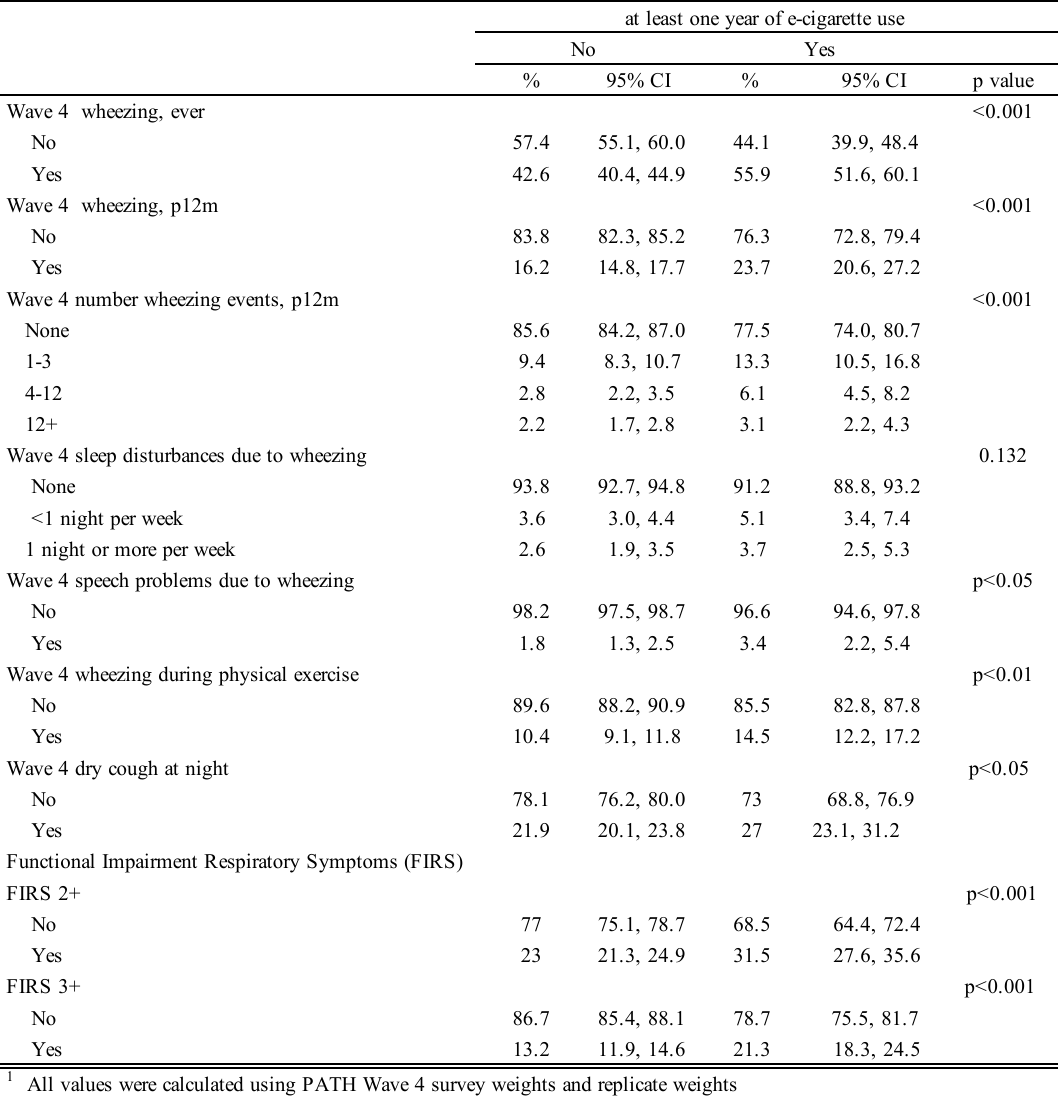

Table A3. Interaction of e-cigarette use duration and respiratory symptoms (FIRS 3+) in relation to incident COPD among ever-smoking adults 40+

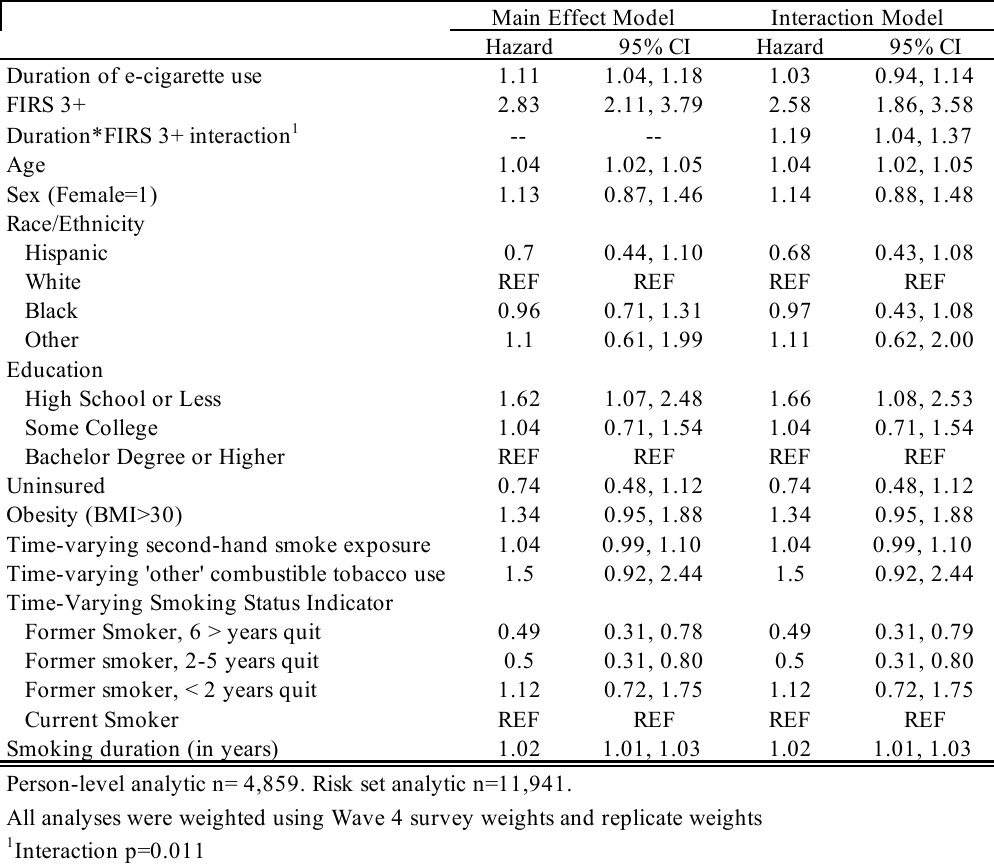

Table A4. E-cigarette use duration and incident COPD among ever-smoking adults 40+, stratified by FIRS 3+

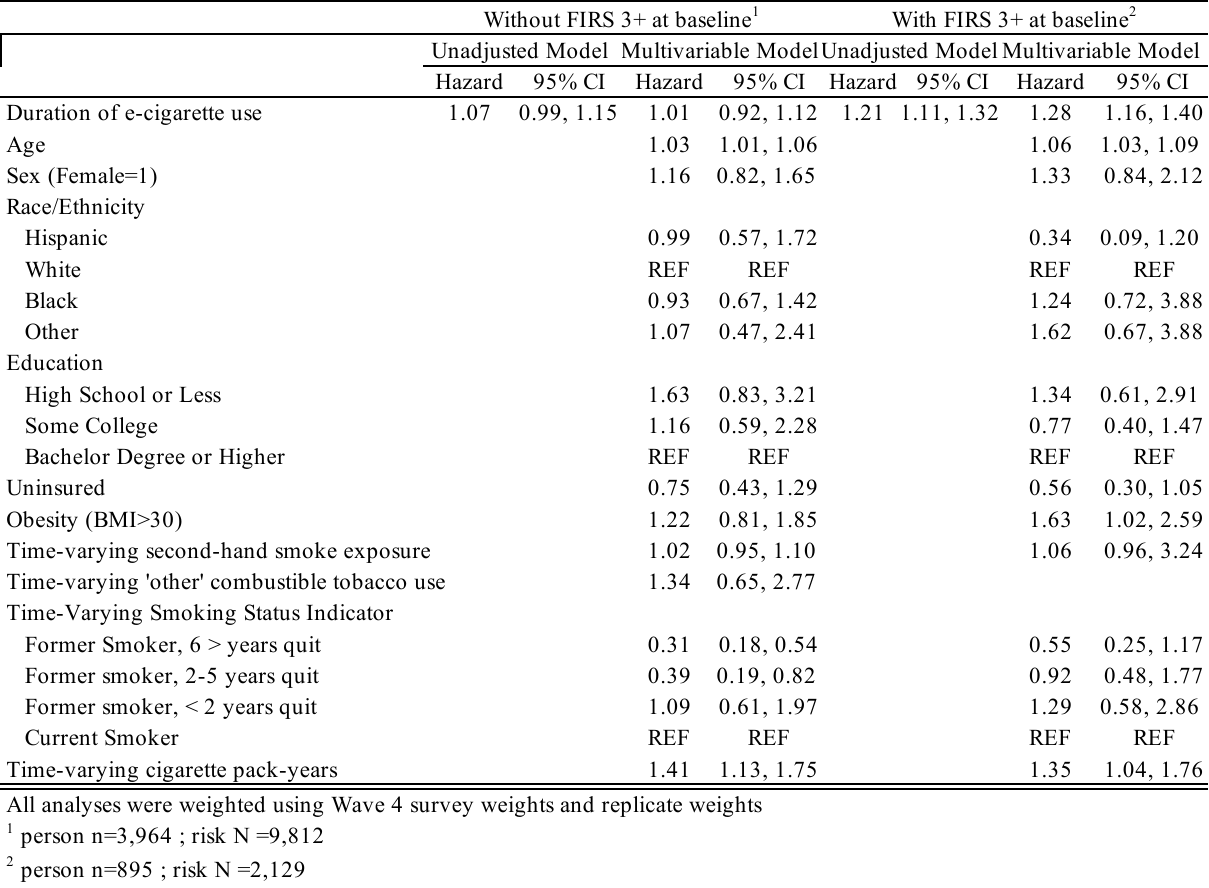

Table A5. E-cigarette use duration and incident COPD among ever-smoking adults 40+, using W4-W7 longitudinal weights

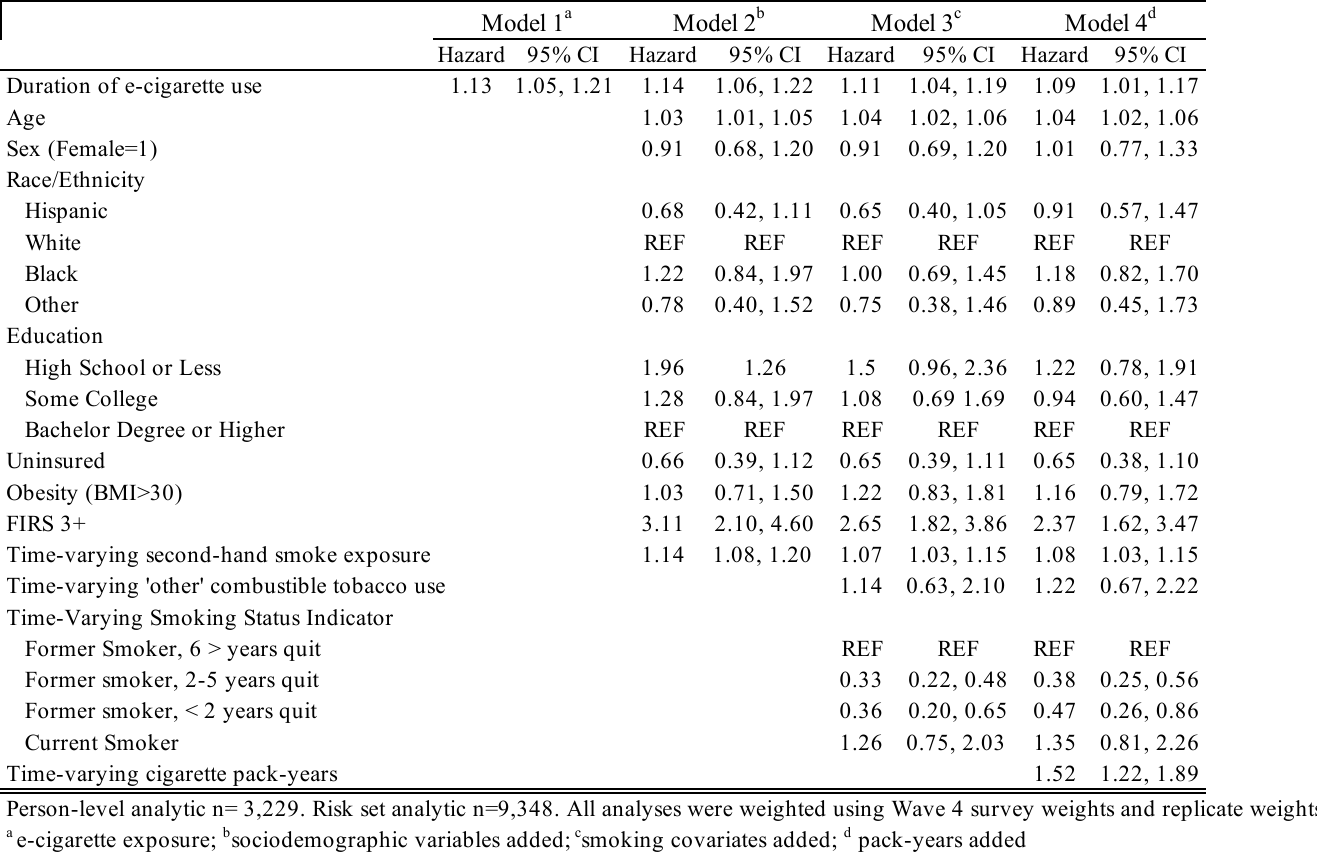

| Table A6. E-cigarette use duration and incident COPD among ever-smoking adults aged 25+ |
| --- |

|  | Model 1^a^ | | Model 2^b^ | | Model 3^c^ | | Model 4^d^ | |
| --- | --- | --- | --- | --- | --- | --- | --- | --- |
|  | Hazard | 95% CI | Hazard | 95% CI | Hazard | 95% CI | Hazard | 95% CI |
| Duration of e-cigarette use | 1.11 | 1.03, 1.19 | 1.16 | 1.10, 1.23 | 1.14 | 1.08, 1.21 | 1.12 | 1.06, 1.19 |
| Age |  |  | 1.04 | 1.03, 1.05 | 1.05 | 1.04, 1.06 | 1.04 | 1.03, 1.06 |
| Sex (Female=1) |  |  | 1.19 | 0.95, 1.49 | 1.18 | 0.94, 1.48 | 1.28 | 1.01, 1.61 |
| Race/Ethnicity |  |  |  |  |  |  |  |  |
| Hispanic |  |  | 0.64 | 0.41, 1.01 | 0.61 | 0.39, 0.94 | 0.78 | 0.51, 1.21 |
| White |  |  | REF | REF | REF | REF | REF | REF |
| Black |  |  | 1.29 | 0.9, 1.73 | 1.03 | 0.78, 1.36 | 1.17 | 0.89, 1.53 |
| Other |  |  | 1.37 | 0.81, 2.32 | 1.31 | 0.77, 2.24 | 1.42 | 0.83, 2.42 |
| Education |  |  |  |  |  |  |  |  |
| High School or Less |  |  | 2.09 | 1.45, 3.02 | 1.65 | 1.13, 2.40 | 1.42 | 0.98, 2.06 |
| Some College |  |  | 1.39 | 0.98, 1.95 | 1.2 | 0.84, 1.72 | 1.08 | 0.75, 1.54 |
| Bachelor Degree or Higher |  |  | REF | REF | REF | REF | REF | REF |
| Uninsured |  |  | 0.95 | 0.64, 1.40 | 0.9 | 0.61, 1.31 | 0.87 | 0.60, 1.27 |
| Obesity (BMI>30) |  |  | 1.19 | 0.88, 1.60 | 1.37 | 1.03, 1.83 | 1.33 | 1.0, 1.77 |
| FIRS 3+ |  |  | 3.21 | 2.39, 4.31 | 2.73 | 2.04, 3.64 | 2.49 | 1.83, 3.39 |
| Time-varying second-hand smoke exposure |  |  | 1.11 | 1.05, 1.15 | 1.05 | 1.0, 1.09 | 1.03 | 0.98, 1.07 |
| Time-varying 'other' combustible tobacco use | |  |  |  | 1.32 | 0.85, 2.04 | 1.38 | 0.90, 2.10 |
| Time-Varying Smoking Status Indicator |  |  |  |  |  |  |  |  |
| Former Smoker, 6 > years quit |  |  |  |  | 0.31 | 0.22, 0.46 | 0.35 | 0.24, 0.50 |
| Former smoker, 2-5 years quit |  |  |  |  | 0.37 | 0.23, 0.58 | 0.46 | 0.30, 0.72 |
| Former smoker, < 2 years quit |  |  |  |  | 0.92 | 0.63, 1.36 | 1.01 | 0.68, 1.49 |
| Current Smoker |  |  |  |  | REF | REF | REF | REF |
| Time-varying cigarette pack-years |  |  |  |  |  |  | 1.37 | 1.17, 1.61 |
| Person-level analytic n= 8,457. Risk set analytic n=20,758. All analyses were weighted using Wave 4 survey weights and replicate weights | | | | | | | | |
| ^a^ e-cigarette exposure; ^b^sociodemographic variables added; ^c^smoking covariates added; ^d^ pack-years added | | | | | | |  |  |

Table A7. E-cigarette use duration and incident COPD among ever-smoking adults 40+ without baseline risk as a covariate

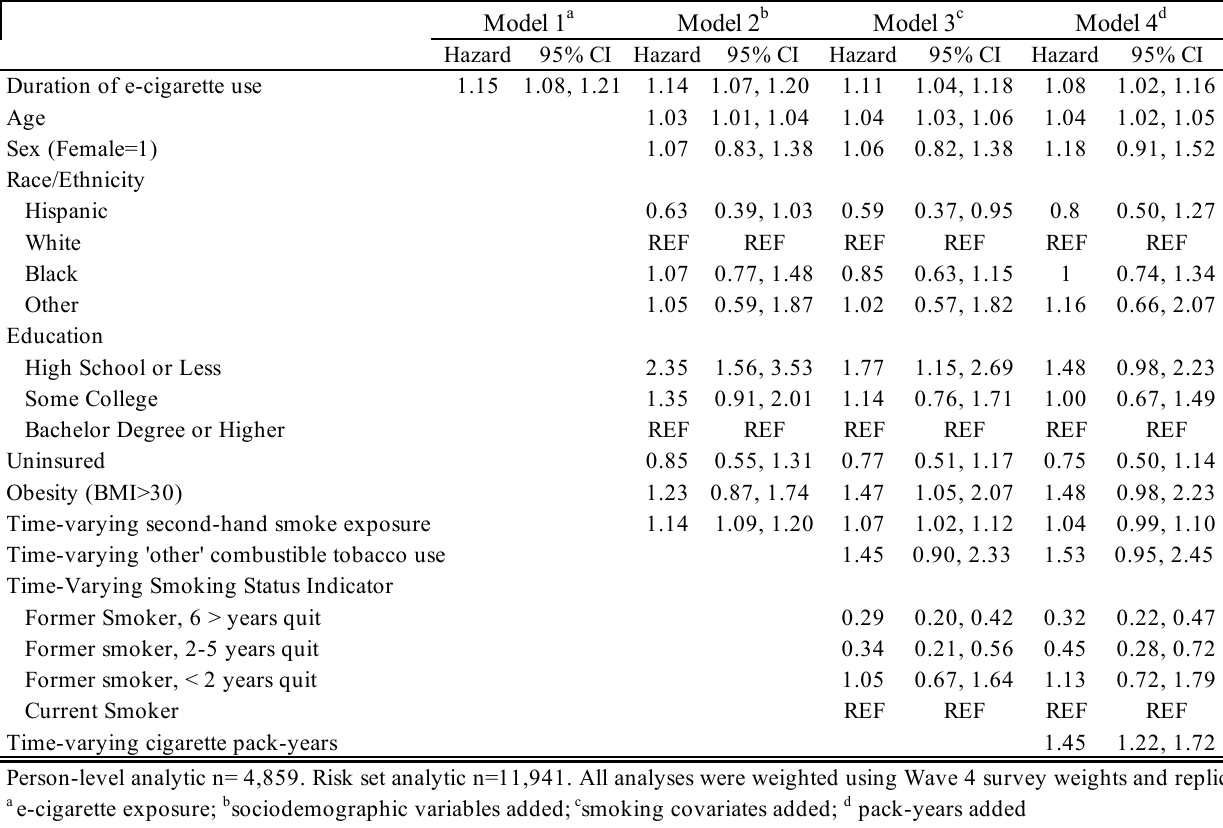

Table A8. Interaction of e-cigarette use duration and cigarette smoking status in relation to COPD among ever-smoking adults 40+

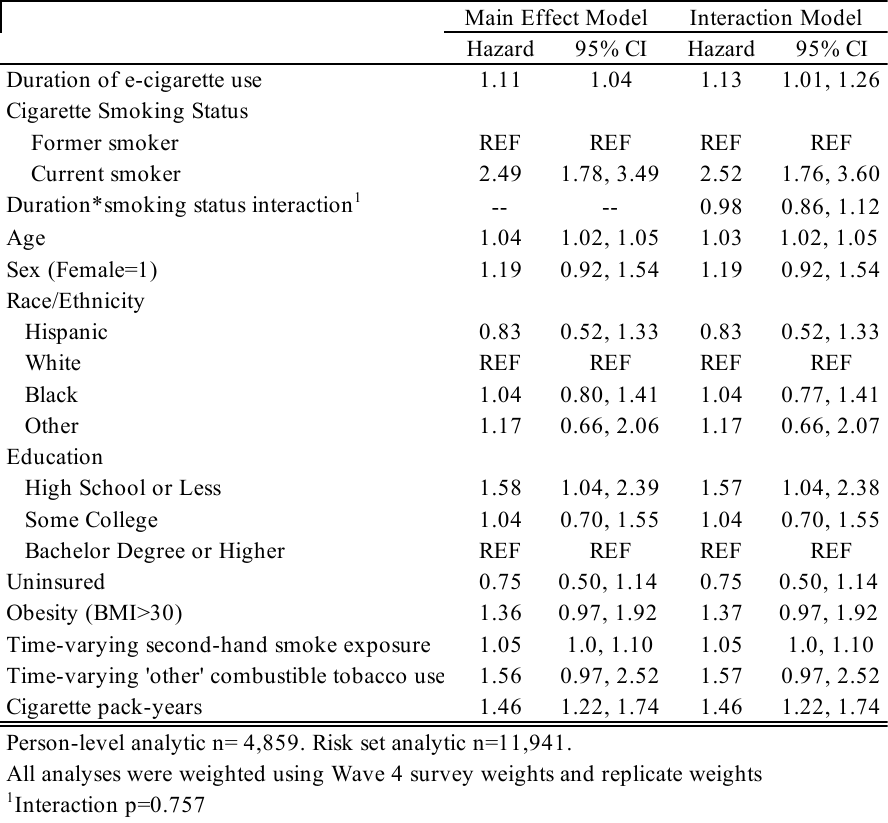

Table A9. Interaction of e-cigarette use duration and cigarette pack-years in relation to COPD among ever-smoking adults 40+

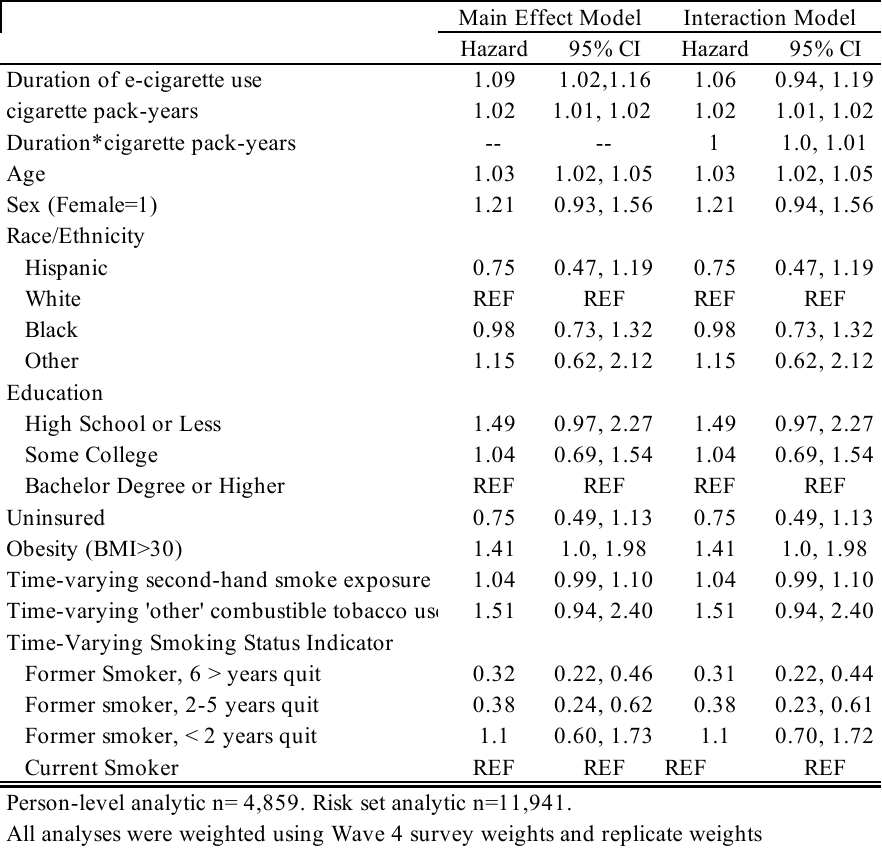

Table A10. Interaction of e-cigarette duration and respiratory symptoms (FIRS 2+) in relation to incident COPD among ever-smoking adults 40+

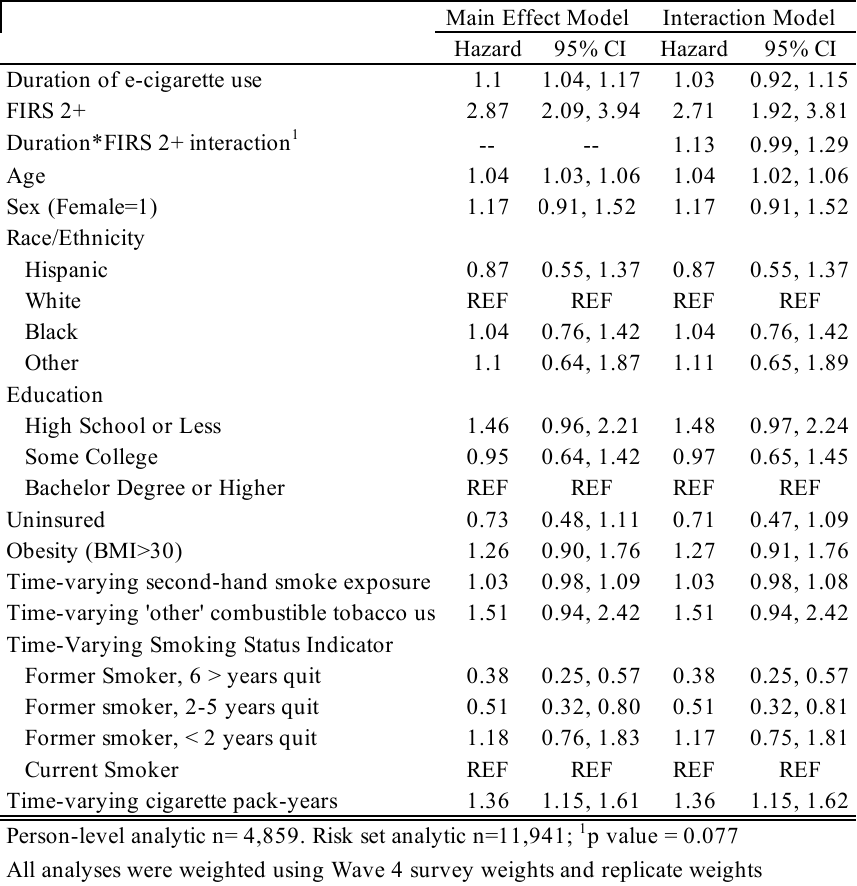

| Table A11. E-cigarette use duration and incident COPD among ever-smoking adults 40+, stratified by FIRS 2+ |
| --- |

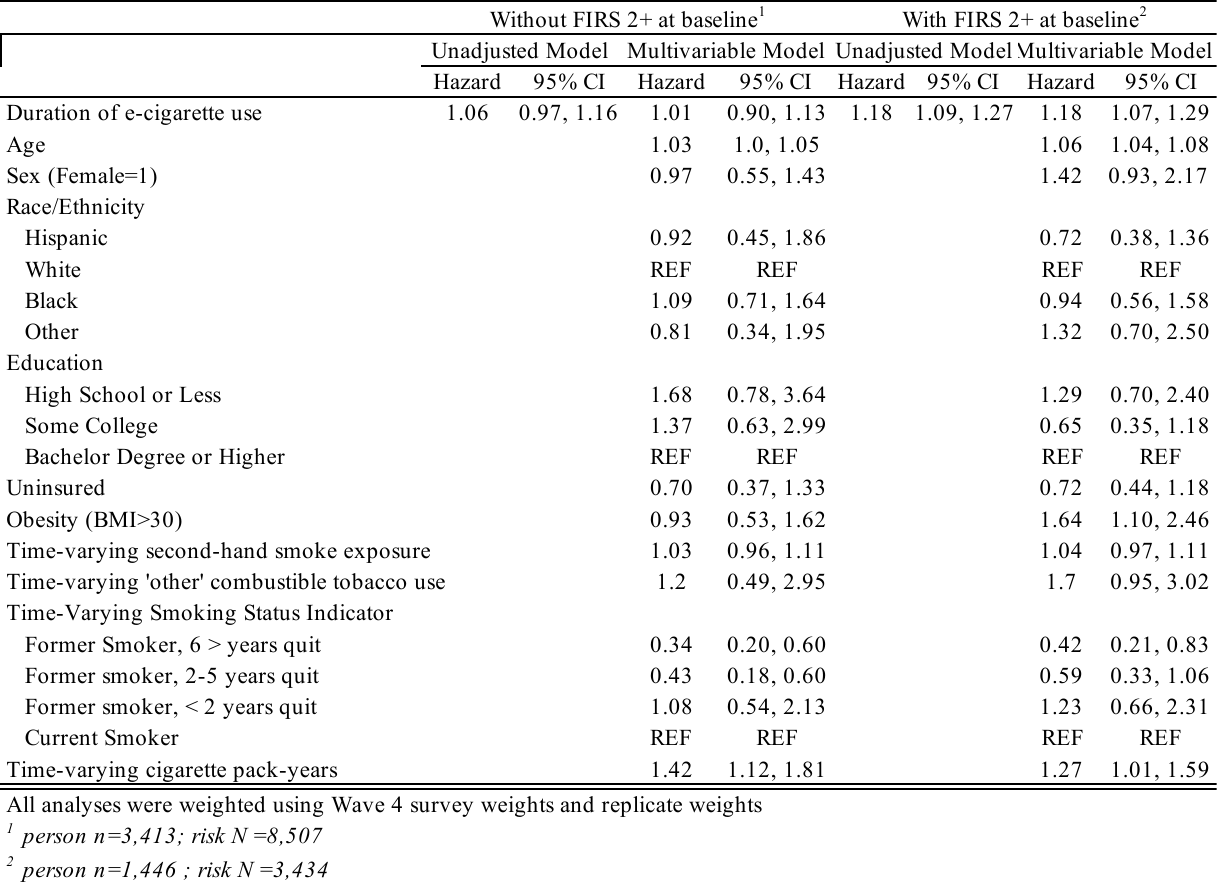
